# Information Technology in Screening and Identifying Unmet Social Needs: A Scoping Review

**DOI:** 10.1101/2024.10.18.24315603

**Authors:** Emre Sezgin, Daniel I Jackson, Samantha Boch, Mattina Davenport, Micah Skeens, Millie Dolce, Bianca Franklin, Lisa K. Militello, Elizabeth Lyman, Kelly Kelleher

## Abstract

**Background:** Social determinants of health (SDOH) affect health outcomes, with unmet social needs like food insecurity, housing instability, and lack of services contributing to poor outcomes, health disparities, and higher healthcare costs. Identifying these needs is vital for improving health and achieving equity. Information technology has emerged as a tool for screening and identifying unmet social needs.

**Objectives:** This scoping review examines how information technology is used to screen and identify unmet social needs among vulnerable populations and its impact on the assessment process.

**Methods:** Following PRISMA-ScR guidelines, we searched databases including MEDLINE, Embase, Scopus, ACM Digital Library, and Web of Science for studies published from 2010 to 2023. Eligible studies used technology to screen for and identify unmet social needs in populations with health and socioeconomic challenges. Data extraction focused on the types of technology, screening processes, and social needs identified.

**Results:** Our findings highlight a limited yet evolving landscape of technological applications. We identified 13 studies using tools like self-assessment surveys, tablet-based systems, and electronic portals. These tools were applied across diverse groups, such as refugees and patients in emergency departments. Innovative approaches, such as chatbots and multi-dimensional risk appraisal systems for older adults, showed potential. However, challenges included single-site studies, small samples, and integration issues with medical records. Effectiveness varied. The effectiveness of these tools in screening for unmet social needs shows mixed outcomes.

**Conclusions:** Information technology plays a pivotal role in improving the identification of unmet social needs. The findings underscore the need for broader, more integrated research to fully understand the impact of technology-based assessments and screening processes for social needs. Future efforts should focus on facilitated screening using technology both within and outside of the visit, ensuring the linkage to appropriate resources and care.

## Introduction

Social determinants of health (SDOH) encompass many elements that profoundly impact individual and population health outcomes.[1] These factors extend beyond healthcare to include environments in which one lives, works, and plays.[2–4] These factors include but are not limited to, economic stability, educational opportunities, social and community contexts, healthcare accessibility, and the physical environment.[5] Unmet social needs (including Health Related Social Needs), such as food insecurity, housing instability, and inadequate access to essential services, are critical aspects of SDOH that are unfortunately widespread.

Empirical evidence has established a strong link between these unmet social needs and adverse health outcomes.[6] For instance, individuals experiencing food insecurity are at greater risk of chronic conditions such as diabetes, heart disease, and obesity.[7] Similarly, housing instability has been associated with an increased risk of mental health disorders, substance use, and infectious diseases.[8–10] Inadequate access to essential services, such as transportation, childcare, and healthcare, can further compound these health challenges.[10–12]

Identifying these unmet social needs is imperative to enhancing overall health and well-being, particularly among historically health-disparate populations experiencing significant health and socioeconomic challenges.[13] Although identifying unmet social needs is recognized as important, integrating screening into routine healthcare remains complex and poorly implemented.[3,14] Unlike social workers and community health workers who specialize in social needs assessment, healthcare providers, such as physicians and advanced practice nurses, often lack the dedicated training, time, and resources necessary to effectively assess the needs during clinical encounters. These providers may find it challenging to incorporate comprehensive screening into their already demanding workloads. This lack of capacity hampers the early identification of unmet social needs, which is crucial for informing patient care and connecting individuals to appropriate support services.[15]

Traditional paper-based screening methods have struggled due to issues with scalability, documentation, resource demands, and inefficient follow-ups.[16] In contrast, the rise of information technology provides opportunities to overcome barriers in screening for unmet social needs. Digital tools, including electronic health records (EHRs), mobile applications and web-based platforms offer scalable and efficient solutions for identifying and detecting unmet social needs.[17,18] Integrating SDOH data into EHRs allows healthcare providers to track changes in patients’ social needs and better understand their circumstances, leading to more personalized care plans.[18] These technologies facilitate the systematic collection, documentation, and analysis of social needs data, enhance patient engagement, and improve referral processes to community resources - important aspects that are also in line with alternative payment or value based care model structures.[19–21] Furthermore, the technology can extend the support beyond clinical settings, to assist patients and families in connecting with essential resources, thereby providing continuous support and improving overall well-being.[22,23] However, current research on information technology-based support and assessment or studies on technology use for screening and detecting unmet social needs is limited.[24] The purpose of this scoping literature review is (1) to examine the extent to which information technology is employed to screen for and identify unmet social needs among populations facing health and socioeconomic challenges, and (2) to investigate the impact of these technologies on screening processes. Finally, we synthesize the findings to provide an overview of the current landscape and implementations, and identify common challenges and limitations. By elucidating the role of technology in identifying unmet social needs, this study seeks to inform future research and clinical practice, ultimately contributing to the development of more effective and equitable healthcare assessment.

## Methods

This scoping review follows the Preferred Reporting Items for Systematic Reviews and Meta-Analyses with the extension for Scoping Reviews (PRISMA-ScR).[25]

### Eligibility Criteria

Within the scope of this review, the studies were selected based on 4 properties (see textbox 1) following Population, Intervention, Comparisons, and Outcomes (PICO) guidelines.[26]

### Textbox 1. PICO guidelines

**Population** (P): Individuals prospectively reported one or more social needs. This includes, but is not limited to, people experiencing social determinants of health issues, socioeconomic factors, health service needs and demands, food insecurity, hunger, or housing insecurity.

**Intervention** (I): Use of technology for screening social needs. This includes various forms of technology such as tablets, iPads, mobile apps, chatbots, kiosks, computers, laptops, and patient portals.

**Comparison** (C): No comparison required but it could involve comparing technological methods of screening for social needs against traditional, non-technological methods, or it could involve comparing different types of technological assessments or interventions against each other.

**Outcome** (O): The outcomes of interest could include the effectiveness of technology in identifying social needs, user satisfaction with the screening tools, integration into clinical workflows, usability, and any challenges encountered during the screening process.

Eligibility criteria of this review included studies (1) prospectively collected study data, (2) reported social need components, (3) demonstrated an information technology-based screening method, and (4) published in English and (5) published between 2010-2023. This review does not include conference abstracts, poster presentations, thesis or dissertations, systematic reviews, literature reviews or meta-analyses, protocol papers, curricula, or publications in non-peer-reviewed journals.

### Search Strategy

We used scientific peer-reviewed academic literature databases including MEDLINE/PubMed, Embase, Scopus, ACM Digital Library, and Web of Science (Figure 1). The search was conducted on July 19, 2023. We extracted peer-reviewed articles (n=2098) using online software, on Covidence software[EL].[27] The researchers [ES, DM, EL, DJ] formed the search queries under 3 groups of keywords for database search: social needs, data collection approach, and technology being used. **Appendix 1** outlines the search query. Additional literature (n=49) was included via searching and reviewing citations from the included papers (n=13).

**Figure 1.**
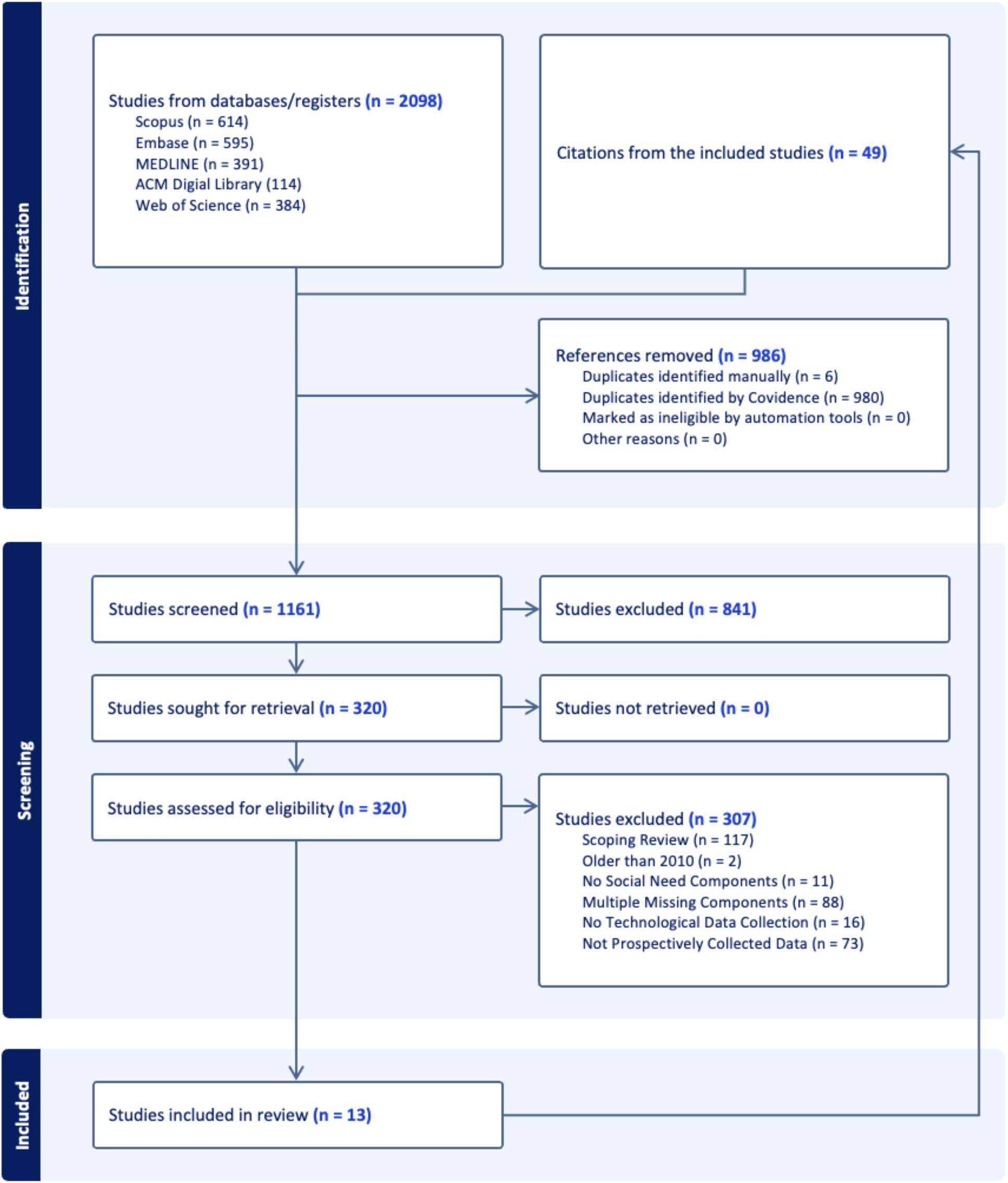
PRISMA-ScR Diagram

### Data Extraction

Articles included in the search were cataloged and systematically reviewed to be included based on the PICO [EL, DIJ, BF, ES]. Duplicate studies were removed automatically (n=980) by Covidence and manually (n=6). Two researchers [DIJ, BF] screened titles and abstracts of the remaining studies (n=1161) and cross-verified and supervised by senior researcher [ES]. Conflicting reviews were resolved by consensus after an open discussion among the team members. Studies were assessed for full eligibility (n=320) and 307 were excluded for various factors such as missing social need components, technological data collection, or were retrospective by design. Thirteen studies were included for full eligibility in the review and were extracted [ES, SB, LM, MS, MD] to an MS Excel file by categorizing the study design, demographics, findings, and other characteristics (**see Appendix 2**).

## Results

### Demographics

The included studies were primarily conducted in the USA, with sample sizes for participants ranging from 13 to 826 (Table 1). Social needs are most frequently assessed in the English language (n=12) and about food insecurity (n=9) and housing instability (n=8). Participants were predominantly female (33.3-63.9%) and White (3.9-86.1%), with notable representation from Black/African American (10.3-63.9%) and Hispanic/Latino/a/X (6.8-52.4%) populations. Tablet computer-based surveys were the most common technology used (n=6), while other technologies like chatbots and software/web apps were less frequently employed (n=2 each). Most studies were published between 2019 and 2023. Most common study outcome reported was the ratio of total patients screened. Participants varied widely in age, race, ethnicity, and socio-economic backgrounds.

**Table 1.** Description of Characteristics of the 13 Included Articles

| Characteristic | Category | # of Articles/Range / Percentage |
| --- | --- | --- |
| Study Locations* | USA | n=9 |
|  | Spain | n=1 |
|  | United Kingdom | n=1 |
|  | Canada | n=2 |
| Sample Sizes of N** | Patients | (13-826) |
|  | Caregivers | (30-505) |
|  | Healthcare Providers | (13-147) |
| Languages** | English | n=12 |
|  | Spanish | n=5 |
|  | Farsi/Dari | n=1 |
| Social Needs Assessed | Food Insecurity | n=9 |
|  | Housing Instability | n=8 |
|  | Healthcare Accessibility | n=7 |
|  | Transportation | n=6 |
|  | Childcare/Education | n=6 |
|  | Financial Stability | n=5 |
|  | Exposure to Violence | n=3 |
| Sex/Gender Identity | %Female | (33.3-63.9%) |
|  | %Male | (35.8-56.7%) |
|  | %Other | (10.0%) |
| Race/Ethnicity of Participants | %White | (3.9-86.1%) |
|  | %Black/African American | (10.3-63.9%) |
|  | %Hispanic/Latino/a/X | (6.8-52.4%) |
|  | %Multiple or other | (0.5-33.4%) |
| Types of Technology | Tablet Survey | n=6 |
|  | Electronic Health Record Portal | n=2 |
|  | Software/Web Apps | n=2 |
|  | Chatbots | n=2 |
|  | Text Messaging | n=1 |
| Study Outcomes | Percentage of Patients Screened | n=10 |
|  | Patient Satisfaction | n=6 |
|  | Patients Referred to Consultation | n=5 |
|  | Number of Healthcare Visits | n=5 |
|  | Patient Health Literacy | n=4 |
|  | Patient Screening Task Load | n=3 |
| Publication Year | 2023 | (23.1%) |
|  | 2022 | (15.4%) |
|  | 2021 | (15.4%) |
|  | 2020 | (15.4%) |
|  | 2019 | (15.4%) |
|  | 2017 | (7.69%) |
|  | 2012 | (7.69%) |

\* Some studies were multisite
\*\* Sample sizes are not mutually exclusive, subsamples derived from a larger sample may overlap

### Types of Technology

In most of the studies, tablet-based computers[28,29] and electronic health record (EHR)-based tools.[30,31] These technologies were used to facilitate the screening, recording, and referral process for unmet social needs within healthcare settings. Web-based systems and electronic portals were also utilized, such as online screener tool[32] or the electronic portal via REDCap (online survey platform) integrated with the 2-1-1 services.[33] Other technologies included cross-platform apps with HIPAA-compliant cloud infrastructure for data collection and analysis,[34] and a software designed for multidimensional risk appraisal in older adults.[35] In addition, text message-based services for social needs screening were used that are linked to REDCap surveys.[36] Similarly, two studies explored text-based screening via innovative approaches like chatbots,[37,38] which leveraged natural language processing to engage users and enhance data collection.

### Unmet Social Needs Assessment

As presented in table 1, the studies examined various social needs. More specifically, these included psychosocial risks for refugees,[28] caregiver needs in pediatric inpatient units,[39] financial needs,[30,32] food insecurity[29,30,39] in primary care settings, and general social needs in diverse patient populations.[32,38] Housing instability and financial strain were also screened in primary care and pediatric settings.[39,40] Furthermore, environmental, psychosocial, and behavioral factors were reported towards reducing cardiovascular risks in African American young adults.[34] During the COVID-19 pandemic, unmet social needs were evaluated in urban emergency departments.[36]

### Research Methods and Approaches

Common methods included mixed-methods and single-arm studies,[29,32,33,38] and quasi-experimental designs.[31,36] Two studies assessed feasibility through simulated clinical workflow implementations.[33,35] Three of the studies reported as quality improvement projects[33,39,40] and a pilot randomized control trial.[28] There were two quality improvement studies used “Plan-Do-Study-Act” cycles to rapid prototyping and adapting the study intervention based on stakeholder (e.g., caregiver, provider, patient) feedback longitudinally.[39,40] In addition, other approaches involved Community-Based Participatory Research (CBPR) and User-Centered Design (UCD) for mobile platforms,[34] within-subjects designs for survey comparisons[37] and validation studies.[30] Studies used a variety of scales for measuring SDOH,[31,37,38] unmet social needs,[28,29,33,35–39] usability,[31,34,37] and cost effectiveness post-intervention,[35] but there were no overlapping measures nor agreement on outcome measures (effect of screening or technology use) observed across the studies.

### Opportunities and challenges

The studies reported a number of opportunities and challenges on the use of technology to identify unmet social needs. In particular cases, patients may express more comfort seeking care services through information technology-based platforms.For example, Ahmad et al. (2012) reported that 72% of participants using a touch-screen self-assessment intended to seek psychosocial counseling compared to 46% in usual care.[28] Commonly identified needs included mental health support, food security, and access to public utilities, with one study noting that 99% of family caregivers received contact information and details for federal and community-based aid programs.[39] Another study indicated participants who self-reported unmet social needs via the information technology-based screening tools had high follow-up rates with local support programs after clinical visits (as high as 86%), leading to sustained patient engagement throughout study procedures.[35] A tablet-based screening approach was reported an increase in screening rates (from 45% to 90%).[40] However, the use of technology did not eliminate the challenges of documentation and management of positive screens, such as missing documents needed to receive community resources.[40] Conversational technologies, such as chatbots, were observed to have impact on the engagement with screening and help with comprehension of social needs related content, particularly among participants with low health literacy.[37,38] Personalization features (language options, multiple modalities, interaction timing, location awareness) were noted as a likely contributor to the adoption of digital screening tools, especially in marginalized groups.[28,32,36,39] However, only text messaging for screening recruitment was deemed infeasible for minority populations.[36] The introduction of electronic methods, like tablet-based systems, resulted in higher social need screening administration rates but introduced new concerns between the personal interactions of patients and providers, leading to a decrease in documentation quality in favor of operational efficiency.[40]

### Key observations

A common observation across the studies was the heterogeneity of the technologies and tools used, such as tablet-based self-assessment systems[28,29] and electronic portals,[33] to facilitate the screening and referral processes for social needs. Several studies focused on specific populations, including Afghan refugees,[28] caregivers in pediatric settings,[39] and patients visiting emergency departments,[33] reporting the technological assessment to diverse demographic groups. Additionally, the majority of studies aimed to evaluate the feasibility and effectiveness of these technological tools in routine healthcare settings, focusing on practical implementation and cost considerations.[29,32,33,35,36,39] Novel approaches and recent technologies are observed, such as chatbots for SDOH screening[37,38] and the integration of multi-dimensional risk appraisal systems.[35] Studies also explored the effectiveness of these approaches in improving social needs-related outcomes associated with quality of life, stress, food insecurity, financial stability, housing stability, and service utilization.[30,31,34,40] Some of the observed outcomes are decreased the risk of cardiovascular disease and improved sleep quality,[34] enhanced the detection of unmet social needs,[30] and created a more equitable system to access community and health services.[31,40] Although survey-style screening tools were the mostly observed in the sample of studies (n=6), software apps and chatbots (n=3) had more study process measures including patient satisfaction and successful resource referrals.

### Limited applications

Common limitations included single-site studies and small sample sizes, affecting generalizability.[28,29,39,40] In addition to that, studies reported the potential self-selection bias and short study durations, impacting the assessment of long-term effects.[37,38] Several studies were missing key demographic features of the study sample. This included the criteria for identifying low digital literacy in patients,[32] socio-economically disadvantaged patients,[30,35] and domain-specific medical conditions.[37] Following that, Walters et al.[35] reported gaps in racial and ethnic community accessibility and increased inequalities in access among certain groups. In terms of technology, Berger-Jenkins et al.[40] noted technology related problems, such as integration challenges with existing medical records systems and documentation difficulties.[41] The inability to track resource map usage or assess successful linkages to community-based organizations was noted as a barrier.[39] With text-based services, it was unknown when the screening completed and if the content is understood well, and noted as some of the potential limitations.[36,37]

## Discussion

In this study, we conducted a scoping review to explore the use of information technology in identifying unmet social needs through screening processes. Our findings reveal a limited but evolving landscape of technological applications, transitioning from basic tablet-based surveys to more sophisticated chatbots. This evolution suggests a potential shift towards facilitating ongoing communication with patients outside of clinical visits, thereby enhancing continuous support and engagement.

Information technology-based screening tools have been useful in screening and identifying unmet social needs by easing the process of screening, leading to an increased number of screening completed. For example, the use of touch-screen self-assessment surveys led to a higher number of requests for psychosocial counseling among participants compared to usual care.[28] Another example was to identify unmet social needs, such as food insecurity and utility assistance and extending to mental health needs, with tablet based or online tools.[29,39] This contributes to the utilization of technology in social services, which has been improving recently, and in consistent with the broad digital health transformation process.[42] The goal is to have improved identification and assessment pipeline through technological integration. However, technology to identify unmet social needs may be required to be a larger effort with organizational or cross-institutional digital transformation to be effective.[43] A Federally Qualified Health Center (FQHC) in Chicago found high technology access and use among patients, with 75% rating the perceived usefulness of patient portals as high.[44] On the other side, technology use was demonstrated by either provider using technology while screening, or patients or individuals completing the screening by using technology themselves. For sensitive topics, individuals may feel more comfortable to self-report without human encounter.[45] That may require further considerations of the utility of the technology regarding the end users. Furthermore, integration of innovative technologies like chatbots may show increased engagement and help comprehension. Particularly among individuals with low health literacy, it can help understand the needs,[37,38] further promoting behavioral interventions and health information communications.[46–48] Innovative and conversational methods may support the process of identification of unmet social needs and interaction to find community services.[48]

The effectiveness of these technological tools in screening for unmet social needs presents mixed outcomes. While tablet-based systems have demonstrated an increase in screening completion rates and higher engagement in seeking services, the long-term impact remains unclear. Furthermore, the findings were limited in demonstrating a sustained long-term impact on identifying unmet social needs. For instance, while an online database helped connect patients with 2-1-1 information specialists within a week of leaving the emergency room,[33] and access to new community resources increased after screening positive for an unmet social need,[29] as well as the rise in social worker consultations,[39] the systemic implementation of such screenings in long term remains challenging.[49–51] Even with these information technology-based assessments, their use has largely been confined to creating electronic resources or registries of patient needs.

Integrating technology with existing systems (e.g., medical records, registries) and documenting social needs are common challenges,[52] which may hinder long-term sustainability.[40] These implementation problems might not be unique, as they have also been reported consistently in a number of studies on adoption digital health and information technologies.[53–55] Additionally, the low recruitment rates and the feasibility issues with the selected technology platforms emphasize the importance of context-specific strategies and technology selections tailored to the target population’s needs and preferences.[35,36] For instance, Pratap et al. study[56] reported that the response and retention to digital tools may show differences depending on the population. Interestingly, as the technology aims to close the gap in accessing services, it could also contribute to widening it. The demographic differences observed in technology adoption (e.g., age, race, literacy) suggest that while technology can enhance access to care, it can also exacerbate existing inequities if not carefully designed and implemented.[35] Therefore, we must consider the barriers specific to different demographic groups to ensure equitable access to health resources. Including diverse teams and stakeholders in the development and deployment of technology to assess social needs is one important way to ensure the wide reach and use of technology in the healthcare setting.[57]

### Comparison with previous literature reviews

Previous reviews focusing on technology use, SDOH and unmet social needs had common themes with our findings.[58–60] Maaß et al.[59] reviewed digital health interventions from a public health perspective. Their work focused on mapping each intervention based on the size of regional application (i.e., proposed, pilot, national, international). Our review expands on the heterogeneity of digital health applications and reports recent new information technology-based screening tools that were adopted for different settings and populations. Yao et al.[58] highlighted the inequities of digital health, with a focus on when health disparities are exacerbated instead of mitigated due to inaccessible information technologies. Similar to our findings, the authors identified a patient’s socioeconomic status as a major contributor to these disparities, as not all have equal access to the technologies. Finally, Craig et al.[60] reported a group of studies under 3 categories in digital health: Policy, Data, and Technology. From a public health perspective, they reported that behavioral-related SDOH assessments and tailored interventions (i.e., weight loss program[61]) may lead to a greater improvement of SDOH outcomes.

### Limitations and future works

The search strategy was confined to the selected databases and only included studies published in English, potentially introducing selection bias. Studies were not regionally restricted to the United States, but due to terminology use, might have resulted in more studies from the U.S. Keywords in our search were limited to SDOH and social needs, not including subcategories or contributing factors to SDOH. The heterogeneity of the assessment approaches and outcomes in the included studies, along with the inclusion criteria, may limit the comprehensiveness and comparability of the findings. The variable quality of evidence, with some studies having methodological limitations further constrained the ability to draw definitive conclusions. The review did not perform a meta-analysis due to the variability in study designs, populations, interventions, and outcomes, which limits the ability to quantify the overall effect of information technology-based assessments.

Future research should include more databases and non-English studies, conduct longitudinal studies, and develop standardized outcome measures to improve the breadth and comparability of reviews. Including the measures to assess how unmet social needs are addressed, and conducting comparative effectiveness and cost-effectiveness analyses can provide more practical insights. Additionally, patient-centered research to understand user experiences and preferences can guide the development of more acceptable and effective screening tools.

### Managerial and practical implications

The findings of this scoping review present several critical implications for key stakeholders involved in addressing unmet social needs through technology. Policymakers must recognize the current scarcity of research on technological interventions for social needs screening and prioritize funding and policies that support the development and evaluation of innovative digital tools, ensuring equitable access to prevent exacerbating existing health disparities. Clinical Informaticians are essential in advancing the evolution of screening technologies from tablets to interactive chatbots, focusing on integration with EHRs and enhancing user experience to facilitate ongoing patient-provider communication outside of clinical visits. Furthermore, social workers, nurses, care coordinators and care teams can leverage these technologies to streamline referral processes and maintain continuous engagement with clients, thereby enhancing support beyond clinical settings. Additionally, healthcare providers may require training and support to effectively implement these tools within clinical workflows, while healthcare administrators may need to allocate resources and facilitate integration with existing systems to ensure the sustainability of technological solutions. Community organizations play a vital role in providing necessary support services, and their collaboration with healthcare institutions is crucial for effective referrals and follow-ups. Engaging patients and caregivers in the design and implementation process ensures that the technologies meet their needs and preferences, enhancing usability and effectiveness. Collaborative efforts among these stakeholders, including technology developers, vendors and researchers, are essential to bridge existing research gaps, optimize the implementation of technological solutions, and ultimately improve health outcomes and achieve greater health equity.

## Conclusion

This work highlights the potential of technology use identifying unmet social needs across diverse populations. The studies reviewed demonstrate the use of various digital tools in enhancing the screening and referral processes for social needs. However, these technologies also present challenges that could potentially exacerbate existing health disparities if not thoughtfully implemented. To fully realize the benefits of information technology-based screening, it is crucial to drive the field toward studying the complete context of the screening process. This includes not only the initial identification of unmet social needs but also the follow-up actions, receipt of services, and the subsequent improvement in health and social outcomes. We suggest more research on comprehensive evaluations that track the continuum from screening to outcomes, ensuring that information technology-based solutions lead to meaningful and sustained improvements in addressing unmet social needs.

## Supporting information

Appendix 1- Search queries

Appendix 2- Literature table

## Acknowledgements

None

## Author contributions

Emre Sezgin led the conceptualization, data curation, formal analysis, investigation, methodology, project administration, and writing – original draft. Daniel I. Jackson contributed to data curation, formal analysis, methodology, project administration, software, resources, visualization, and writing – review & editing. Samantha Boch, Mattina Davenport, Micah Skeens, Dolce Millie, and Lisa K. Militello were involved in investigation, validation, and writing – review & editing. Bianca Franklin provided support with data curation, resources, and writing – review & editing. Elizabeth Lyman contributed to data curation, formal analysis, methodology, software, resources, and writing – review & editing. Kelly Kelleher played a key role in investigation, supervision, and writing – review & editing. All authors reviewed and approved the final manuscript.

## Data availability

The data used in this article are available in the supplementary material.

