## Appendix 1- Search queries for "Information Technology in Screening and Identifying Unmet Social Needs: A Scoping Review"

### Embase

595 results

Searched 7-19-23

#### Topic 1:

('social determinants of health'/de OR 'social determinant*':ti,ab OR 'social factors determining health':ti,ab OR 'social health determinant*':ti,ab OR 'sdoh':ti,ab OR 'determinants of health':ti,ab OR 'socioeconomic determinant*':ti,ab OR 'social needs'/de OR 'social needs':ti,ab OR 'social risk*':ti,ab)

AND

#### Topic 2:

(checklist*:ti,ab OR 'mass screening'/de OR screen*:ti,ab OR 'needs assessment'/de OR 'social needs assessment':ti,ab OR 'questionnaire'/de OR 'survey*':ti,ab OR 'questionnaire*':ti,ab)

AND

#### Topic 3

('automated conversation pathway*':ti,ab OR avatar*:ti,ab OR 'chatbot*':ti,ab OR ((conversation* OR 'interactive voice' OR 'virtual' OR 'voice' OR 'voice activated' OR 'voice assistant*' OR 'voice controlled' OR 'voice enabled' OR 'voice initiated' OR 'voice interactive') NEAR/3 (app OR apps OR application* OR agent* OR agents OR assistant* OR device* OR interface* OR system* OR technolog*)) OR 'dialogue system*':ti,ab OR 'digital personal assistant*':ti,ab OR 'integrated smartphone system*':ti,ab OR 'intelligent personal assistant*':ti,ab OR (mobile NEAR/3 (app OR apps OR application* OR assistant*)) OR 'personal digital assistant*':ti,ab OR 'short message service':ti,ab OR 'smart speaker*':ti,ab OR (smartphone NEAR/3 (application* OR app OR apps)) OR 'speech recognition':ti,ab OR 'speech to text':ti,ab OR 'telephone linked care technolog*':ti,ab OR 'text messag*':ti,ab OR 'sms':ti,ab OR 'text to speech':ti,ab OR 'user computer interface*':ti,ab OR 'voicebot':ti,ab OR 'patient portal*':ti,ab OR 'electronic medical record*':ti,ab OR 'electronic health record*':ti,ab OR 'emr':ti,ab OR 'ehr':ti,ab OR 'computer*':ti,ab)

#### Ovid Medline

391 results

Searched 7-19-23

("Social Determinants of Health"/ OR "social determinant*".mp OR "social factors determining health".mp OR "social health determinant*".mp OR "sdoh".mp OR "determinants of health".mp OR "socioeconomic determinant*".mp OR "social needs".mp OR "social risk*".mp) AND (checklist*.mp OR "Mass Screening"/ OR screen*.mp OR "needs assessment"/de OR "social needs assessment".mp OR "Surveys and Questionnaires"/ OR "survey*".mp OR "questionnaire*".mp) AND ("automated conversation pathway*".mp OR avatar*.mp OR "chatbot*".mp OR ((conversation* OR "interactive voice" OR "virtual" OR "voice" OR "voice activated" OR "voice assistant*" OR "voice controlled" OR "voice enabled" OR "voice initiated" OR "voice interactive") ADJ3 (app OR apps OR application* OR agent* OR agents OR assistant* OR device* OR interface* OR system* OR technolog*)) OR "dialogue system*".mp OR "digital personal assistant*".mp OR "integrated smartphone system*".mp OR "intelligent personal assistant*".mp OR (mobile ADJ3 (app OR apps OR application* OR assistant*)) OR "personal digital assistant*".mp OR "short message service".mp OR "smart speaker*".mp OR (smartphone ADJ3 (application* OR app OR apps)) OR "speech recognition".mp OR "speech to text".mp OR "telephone linked care technolog*".mp OR "text messag*".mp OR "sms".mp OR "text to speech".mp OR "user computer interface*".mp OR "voicebot".mp OR 'patient portal*'.mp OR 'electronic medical record*'.mp OR 'electronic health record*'.mp OR 'emr'.mp OR 'ehr'.mp OR 'computer*'.mp)

#### Web of Science

384 results

Searched 7-19-23

TS=(("social determinant*" OR "social factors determining health" OR "social health determinant*" OR "sdoh" OR "determinants of health" OR "socioeconomic determinant*" OR "social needs" OR "social risk*") AND

(checklist* OR "Mass Screening" OR screen* OR "needs assessment"de OR "social needs assessment" OR "Surveys and Questionnaires" OR "survey*" OR "questionnaire*") AND

("automated conversation pathway*" OR avatar* OR "chatbot*" OR ((conversation* OR "interactive voice" OR "virtual" OR "voice" OR "voice activated" OR "voice assistant*" OR "voice controlled" OR "voice enabled" OR "voice initiated" OR "voice interactive") NEAR/3 (app OR apps OR application* OR agent* OR agents OR assistant* OR device* OR interface* OR system* OR technolog*)) OR "dialogue system*" OR "digital personal assistant*" OR "integrated smartphone system*" OR "intelligent personal assistant*" OR (mobile NEAR/3 (app OR apps OR application* OR assistant*)) OR "personal digital assistant*" OR "short message service" OR "smart speaker*" OR (smartphone NEAR/3 (application* OR app OR apps)) OR "speech recognition" OR "speech to text" OR "telephone linked care technolog*" OR "text messag*" OR "sms" OR "text to speech" OR "user computer interface*" OR "voicebot" OR "patient portal*" OR "electronic medical record*" OR "electronic health record*" OR "emr" OR "ehr" OR "computer*"))

#### Scopus

614 results

Searched 7-19-23

TITLE-ABS-KEY (("social determinant*" OR "social factors determining health" OR "social health determinant*" OR "sdoh" OR "determinants of health" OR "socioeconomic determinant*" OR "social needs" OR "social risk*") AND

(checklist* OR "Mass Screening" OR screen* OR "needs assessment" OR "social needs assessment" OR "Surveys and Questionnaires" OR "survey*" OR "questionnaire*") AND

("automated conversation pathway*" OR avatar* OR "chatbot*" OR ((conversation* OR "interactive voice" OR "virtual" OR "voice" OR "voice activated" OR "voice assistant*" OR "voice controlled" OR "voice enabled" OR "voice initiated" OR "voice interactive") NEAR/3 (app OR apps OR application* OR agent* OR agents OR assistant* OR device* OR interface* OR system* OR technolog*)) OR "dialogue system*" OR "digital personal assistant*" OR "integrated smartphone system*" OR "intelligent personal assistant*" OR (mobile NEAR/3 (app OR apps OR application* OR assistant*)) OR "personal digital assistant*" OR "short message service" OR "smart speaker*" OR (smartphone NEAR/3 (application* OR app OR apps)) OR "speech recognition" OR "speech to text" OR "telephone linked care technolog*" OR "text messag*" OR "sms" OR "text to speech" OR "user computer interface*" OR "voicebot" OR "patient portal*" OR "electronic medical record*" OR "electronic health record*" OR "emr" OR "ehr" OR "computer*"))

#### ACM Digital Library- ACM Full-Text Collection

114 results

Searched 7-19-23

(("Title:("social determinant*" OR "social factors determining health" OR "social health determinant*" OR "sdoh" OR "determinants of health" OR "socioeconomic determinant*" OR "social needs" OR "social risk*") OR (Abstract:("social determinant*" OR "social factors determining health" OR "social health determinant*" OR "sdoh" OR "determinants of health" OR "socioeconomic determinant*" OR "social needs" OR "social risk*")))
